# Clustering of characteristics associated with level of pregnancy intention: a latent class analysis of an urban cross-sectional sample in the Netherlands

**DOI:** 10.64898/2025.12.22.25342800

**Authors:** Merel Sprenger, Matty Crone, Joyce M. Molenaar, M. Nienke Slagboom, Jessica C. Kiefte-de Jong

**Affiliations:** Health Campus The Hague/Department of Public Health and Primary Care, Leiden University Medical Center, The Hague, The Netherlands; Department of Health Promotion, Research Insitutes CAPHRI and NUTRIM, Maastricht University, Maastricht, The Netherlands; Department for Population Health and Health Services Research, National Institute for Public Health and the Environment, Bilthoven, the Netherlands

**Keywords:** pregnancy intention, social determinants of health, latent class analysis, unintended pregnancy, unplanned pregnancy

## Abstract

**Background:** Despite evidence that pregnancy intentions are complex, unintended pregnancy remains studied using binary measures and few studies have examined combinations of factors contributing to pregnancy intention. This study aimed to identify groups of pregnant people with similar combinations of characteristics and exposures, and study the association between these groups and level of pregnancy intention.

**Methods:** This study, part of a population-based cohort study of pregnant people and partners (RISE UP study), uses cross-sectional surveys linked to routine data. Latent class analysis identified groups of pregnant people in The Hague distinguished by shared socioeconomic and reproductive characteristics and positive and negative exposures. Linear regression assessed the association between class membership and level of pregnancy intention, adjusted for recruitment location, gestational age at survey, and survey year.

**Results:** In the final sample of 560 pregnant people, four classes were identified. Two reflected general stability, differing by gravidity: *high stability, multigravida* and *high stability, primigravida*. Two reflected greater socioeconomic adversity and fewer positive exposures, differing by presence of negative exposures: *partial stability* and *cumulative adversity*. Compared to the *high stability, multigravida* class, pregnancy intention was significantly lower in the *partial stability* class (adjusted beta = -0.71 95%CI (-1.31 – -0.11)) and the *cumulative adversity* class (adjusted beta = -1.81 95%CI (-2.31 – -1.30)).

**Discussion:** Cumulative adversity and partial stability were associated with lower pregnancy intention, supporting suggestions that pregnancy intention may indicate underlying systemic inequalities. Policy and care providers, including midwives, should address these inequalities and tailor support to individual needs.

## Introduction

Pregnancy intention has long been measured as a binary concept: planned or unplanned, wanted or unwanted, intended or unintended. Within this binary, unintended pregnancies have been associated with higher odds of depression during and after pregnancy, interpersonal violence, preterm birth, and low birth weight.^1^ However, pregnancy intention is influenced by many different aspects including someone’s plans, perceptions and feelings towards pregnancy, and binary measures fail to reflect this complexity.^2-5^ Rather than relying on the binary, quantitative measures can allow for variation across multiple dimensions, resulting in a range of pregnancy intentions from very unintended to more ambivalent to very intended.^6^

Moreover, having lower pregnancy intentions, i.e., a more unintended pregnancy, does not inevitably lead to adverse medical or (psycho)social outcomes, since pregnancy occurs within a broader context and outcomes depend on a combination of characteristics and exposures.^7^ By considering combinations of characteristics and exposures, rather than isolating singular characteristics, we could move towards identifying groups of people with shared characteristics and exposures, who might also exhibit distinct levels of pregnancy intention. This is crucial for improved recognition of different pregnancy intentions and, more specifically, people who might need more or different types of care or support.

Singular characteristics associated with more unintended pregnancies include being single or in a non-cohabiting relationship, younger or older (i.e., age<30 and age>39), having a lower income or education level, having lower social support, experiencing adverse childhood events, periconceptional substance use and psychological distress.^8-16^ Studies reported mixed results regarding the association between parity and unintended pregnancy.^8,17^ A recent study, using a binary measure, investigated clustering of sociodemographic, mental and physical health, and reproductive characteristics in a subsample of women with an unplanned pregnancy.^18^ Four clusters were found: the first cluster was older, with higher income and education level, and history of depression. The second was more often married, multipara, Islamic, with a migration background, later sexual debut, and lower self-esteem. People in the third and fourth cluster were mainly younger, single, with lower income and education level, more childhood trauma and lower self-esteem, either with (third) or without migration background (fourth).^18^ As the clusters were distinguished in a subsample of women with an unintended pregnancy, these findings provide valuable insight into which characteristics cluster among this group, but not which combinations of characteristics are associated with pregnancy intention. Moreover, pregnancy intention was measured as a binary variable rather than on a continuum allowing for a variation in intentions.

Arguing for a broader, contextual, understanding of unintended pregnancy and using a continuous measure, this study aims to first identify groups of pregnant people with shared combinations of socioeconomic and reproductive characteristics and positive and negative exposures, and second, to study the association between group membership and pregnancy intentions.

## Methods

### Study design and population

This study is based on cross-sectional surveys linked to routine data and was embedded in a larger mixed methods population-based cohort study called the RISE UP (RISk and rEsilience in Unintended Pregnancy) study. The survey study was exempted from review by the Medical Research Ethics Committee Leiden Den Haag Delft under reference number N21.127, and was supported by ZonMw under Grant numbers 554002006 and 554001004. Survey data was collected from January 2022 to September 2024 and linked to non-public microdata from Statistics Netherlands (SN).^19^ To reduce information bias, four experts-by-experience, three women and one man, who had experienced a pregnancy that they perceived as more unintended participated in think aloud interviews, commenting on the burden (e.g., length, confronting questions, and understandability) of the survey, recruitment, and compensation for study participation. Additionally, before and during data collection, survey and study design was discussed with an expert team including midwives, a general practitioner, and social workers. To limit survey length, some survey items were omitted if they were present in SN microdata.

Individuals residing in The Hague, the Netherlands, who were pregnant were eligible for inclusion, as were their (ex-)partners, defined as individuals with whom they were in a romantic relationship or had had sexual contact. To reduce selection bias, recruitment took place across multiple settings, with additional efforts in locations where recruitment was initially lower: researchers, research assistants, or health care professionals informed participants at nine Youth and Family Centres (when participants got whooping cough vaccination in pregnancy), 20 midwifery practices, an obstetrics ward, several general practices, and two locations providing options counselling or through advertisements on social media and on an online pregnancy platform, resulting in a convenience sample. Participants were provided with the information letter and video and could ask any questions to the person informing them or the coordinating researcher. After deciding to participate, they gave informed consent, including whether they consented to link their survey results to SN microdata (86% consented, of them 90% were successfully linked based on date of birth, sex at birth, and address). Participants could choose to complete the survey online, in hard copy, or in an interview. All options were available in Dutch and English, hard copy surveys were available in Turkish, Arabic, Mandarin, Polish, and Bulgarian. Upon completing the 15-minute survey, they received a voucher worth €10.

### Measures

Information bias was mitigated by using validated instruments where available. Socioeconomic characteristics included age, origin, religion, marital status, single household, education level, full-time employment, perceived income, household income, and home ownership. Reproductive characteristics included gestational age at positive test, gravidity, previous miscarriage or abortion, and age at sexual debut. Exposures were selected based on factors associated with vulnerability in pregnancy based on Van der Meer et al.’s model, earlier studies,^7,20-24^ and on factors associated with unintended pregnancy.^8-11^ Negative exposures included worries, recent abuse, adverse life events, substance use, and high GP and hospital health expenditures; positive exposures included perceived wellbeing, resilience, and support.^6,25-31^ At the end of the survey, respondents were asked if they participated in any interventions, if they needed help or support and if they consented to being contacted by us about this for referral to suitable support.

To measure pregnancy intention, we used the Dutch Adaptation of the London Measure of Unplanned Pregnancy (DA-LMUP), a reliable and valid modified measure inspired by the structure of the London Measure of Unplanned Pregnancy, resulting in a scale from 0 to 12, with a score of 0 indicating that a pregnancy is very unintended and a score of 12 indicating that a pregnancy is very intended.^6,31^

Categorisation of numerical variables was done based on the distribution. If the distribution was normal, cut-offs were based on the mean and standard deviation (SD). If the distribution was skewed, cut-offs were based on the interquartile range (IQR) or the median. Some variable categories were merged to better suit analyses. An overview of measures, definitions, categorisation, and data sources can be found in Supplementary Table 1. The survey can be found on the RISE UP study OSF project, doi: 10.17605/OSF.IO/63HBE.

### Statistical analysis

All analyses were performed in R version 4.4.3. Data were cleaned, descriptive statistics were computed (n/%, median/IQR, and mean/SD). Bias due to missing survey data (range: 0.2% to 5.7%) or missing SN microdata (range: 9.4% to 20.3%) due to unsuccessful links was addressed through Multiple Imputation using classification and regression trees (CART), yielding five imputed datasets.^32^ Since our analysis cannot be run with pooled datasets, one imputed dataset was randomly selected for subsequent analyses. Given potential differences regarding randomly selecting one imputed dataset, sensitivity analyses were performed with a different imputed dataset, yielding comparable results.

To identify groups of people with different combinations of characteristics and exposures, we performed a latent class analysis (LCA). This is a data-driven method of analysis guided by data with the goal of organising the individuals in the data into unobserved subgroups based on their highest posterior probability: latent classes.^33^ Each of these classes is then characterised by its proportion across all categories of each variable included in the analysis. Using the poLCA package,^34^ we assessed different latent class models including all socioeconomic and reproductive characteristics, positive and negative exposures, and excluding pregnancy intention. We did not have any assumptions on the ideal number of classes. Due to our sample size, statistical fit was evaluated with the adjusted Bayesian Information Criterion (aBIC) where a lower statistic suggests a more appropriate model fit.^35^ In addition, each model’s entropy was considered, indicating the level of certainty of class assignment through a score between 0 (least certain) and 1 (most certain).^36^ Based on the aBIC and entropy, we compared three models’ item-response probabilities and subsequently selected the model that was most interpretable and informative. Finally, the association between class membership and pregnancy intention was investigated through a linear regression analysis, adjusting for recruitment location (i.e., Centre for Youth and Family, primary care midwife, online, other), gestational age at survey, and survey year to account for potential differences in recruitment context over time. A p-value of <0.05 was considered statistically significant.

## Results

In total, 935 individuals (765 pregnant people, 170 partners) started the survey. Sufficient data was collected for 800 participants (655 pregnant people, 145 partners) and approximately 85% of those individuals consented to SN link. Descriptive statistics of the RISE UP participants, stratified by consent for the SN link, are presented in Supplementary Table 2. Compared to those who consented, participants without consent tended to be younger, more frequently religious, married, and with a migration background. They also reported a lower perceived income, later sexual debut, less alcohol use, and lower levels of resilience and support. In addition, their DA-LMUP scores showed fewer instances of the maximum score of 12. Table 1 presents descriptive characteristics of pregnant people (n = 560) and partners (n = 125) who consented to SN link. Not enough partners participated to perform our planned analyses for this group.

**Table 1.** Characteristics of pregnant people and partners, n (%) unless stated otherwise.

| Measure | Pregnant people<br>(n = 560) | Partners<br>(n=125) |
| --- | --- | --- |
| **Socioeconomic characteristics** |  |  |
| **Age – mean (SD)** | 31.82 (4.50) | 33.04 (5.38) |
| Under 25 | 35 (6) | 5 (4) |
| 25 to 29 | 125 (22) | 25 (20) |
| 30 to 34 | 245 (44) | 55 (44) |
| Over 34 | 155 (28) | 40 (32) |
| **Gender** |  |  |
| Man | 0 (0) | 115 (92) |
| Woman | 560 (100) | 5 (4) |
| Other | 5 (1) | 0 (0) |
| **Origin** |  |  |
| The Netherlands | 335 (60) | 85 (68) |
| Abroad: one or both parents | 100 (18) | 20 (16) |
| Abroad: self | 130 (23) | 20 (16) |
| **Religion** |  |  |
| No religion | 370 (66) | 85 (68) |
| Christian/Jewish | 105 (19) | 20 (16) |
| Muslim | 60 (11) | 10 (8) |
| Other | 25 (4) | 5 (4) |
| **Relationship status** |  |  |
| Single | 30 (5) | 0 (0) |
| Non-cohabiting relationship | 20 (4) | 0 (0) |
| Cohabiting relationship | 250 (45) | 50 (40) |
| Married | 260 (46) | 70 (56) |
| **Single household** |  |  |
| No | 460 (82) | 110 (88) |
| Yes | 100 (18) | 10 (8) |
| **Education level** |  |  |
| Low | 25 (4) | 5 (4) |
| Middle | 140 (25) | 35 (28) |
| High | 395 (71) | 80 (64) |
| **Employment** |  |  |
| None | 75 (13) | 5 (4) |
| Part-time | 210 (38) | 25 (20) |
| Full-time | 275 (49) | 90 (72) |
| **Perceived income** |  |  |
| (Very) difficult | 30 (5) | 10 (8) |
| Coping | 120 (21) | 30 (24) |
| Comfortable | 410 (73) | 85 (68) |
| **Income percentile – median (IQR)** | 79 (55-90) | 84 (67-92) |
| Low | 145 (26) | 35 (28) |
| Middle | 285 (51) | 60 (48) |
| High | 130 (23) | 30 (24) |
| **Home ownership** |  |  |
| No | 245 (44) | 45 (36) |
| Yes | 315 (56) | 75 (60) |
| **Reproductive characteristics** |  |  |
| **Gestational age at positive test – median (IQR)** | 4 (4-5) | 4 (3-5.5) |
| Median and under | 355 (63) | 80 (64) |
| Over median | 205 (37) | 45 (36) |
| **Gestational age at survey – median (IQR)** | 24 (15-27) | 25 (14-28) |
| **Gravidity** |  |  |
| Primigravida | 250 (45) | 70 (56) |
| Multigravida | 315 (56) | 50 (40) |
| **Previous miscarriage** |  |  |
| No | 435 (78) | 95 (76) |
| Yes | 125 (22) | 25 (20) |
| **Previous abortion** |  |  |
| No | 500 (89) | 120 (96) |
| Yes | 60 (11) | 5 (4) |
| **Age at sexual debut – median (IQR)** | 17 (16-19) | 18 (15.5-20.0) |
| Low | 225 (40) | 30 (24) |
| Middle | 210 (38) | 65 (52) |
| High | 125 (22) | 25 (20) |
| **Negative exposures** |  |  |
| **Worries – median (IQR) average score** | 1.38 (1.13-1.88) | 1.5 (1.19-1.75) |
| Low | 150 (27) | 30 (24) |
| Middle | 310 (55) | 65 (52) |
| High | 100 (18) | 25 (20) |
| **Recent abuse** |  |  |
| No | 540 (96) | 115 (92) |
| Yes | 20 (4) | 5 (4) |
| **Adverse life events – median (IQR)** | 3 (1-4) | 3 (1-4) |
| Zero | 25 (4) | 5 (4) |
| One to three | 360 (64) | 65 (52) |
| Four or more | 175 (31) | 50 (40) |
| **Alcohol use** |  |  |
| Never | 140 (25) | 30 (24) |
| Stopped during pregnancy | 395 (71) | 20 (16) |
| Use during pregnancy | 30 (5) | 75 (60) |
| **Smoking** |  |  |
| Never | 450 (80) | 100 (80) |
| Stopped during pregnancy | 75 (13) | 5 (4) |
| Use during pregnancy | 35 (6) | 15 (12) |
| **Drug use** |  |  |
| Never | 475 (85) | 95 (76) |
| Before or during pregnancy | 85 (15) | 25 (20) |
| **GP expenditures (€) – median (IQR)** | 146.5 (102.7-235.5) | 129.6 (100.4-187.8) |
| Low | 140 (25) | 30 (24) |
| Middle | 280 (50) | 65 (52) |
| High | 140 (25) | 30 (24) |
| **Hospital expenditures (€) – median (IQR)** | 81.05 (0-617.29) | 0 (0-188.4) |
| No costs | 220 (39) | 75 (60) |
| Lower | 170 (30) | 25 (20) |
| Higher | 170 (30) | 25 (20) |
| **Positive exposures** |  |  |
| **Support – median (IQR)** | 27 (24-29) | 25 (23-28) |
| Low | 170 (30) | 40 (32) |
| Middle | 260 (46) | 60 (48) |
| High | 130 (23) | 25 (20) |
| **Wellbeing – mean (SD)** | 24.46 (3.59) | 22.26 (2.87) |
| Low | 105 (19) | 20 (16) |
| Middle | 355 (63) | 85 (68) |
| High | 100 (18) | 20 (16) |
| **Resilience – median (IQR)** | 7 (6-8) | 7 (6-8) |
| Lower | 225 (40) | 45 (36) |
| Higher | 335 (60) | 80 (64) |
| **Recruitment characteristics** |  |  |
| **Recruitment location** |  |  |
| Youth and Family Centre | 225 (40) | 30 (24) |
| Primary care midwife | 140 (25) | 50 (40) |
| Online | 130 (23) | 10 (8) |
| Other | 65 (12) | 30 (24) |
| **Survey year** |  |  |
| 2022 | 120 (21) | 30 (24) |
| 2023 | 265 (47) | 50 (40) |
| 2024/2025 | 180 (32) | 45 (36) |
| **Outcome** |  |  |
| **Pregnancy intention – median (IQR)** | 11 (10-12) | 10 (9-11) |

Note: table reports imputed data. Numbers rounded to five and percentages presented as integer numbers in accordance with Statistics Netherlands guidelines to prevent disclosure of information about individuals. Hence, percentages may not sum to exactly 100.

### Latent class analysis

Table 2 presents the results of the latent class analysis. Based on the fit indices (Supplementary Table 3), we compared models with four, five, and six classes. A four-class model was found to be most suitable as this was the class where the aBIC showed a clear improvement before levelling off, the entropy (0.93) was highest, and it had the best interpretability.

**Table 2.** Class proportions of the final four-class model for pregnant people.

| Class | High stability, multigravida | High stability, primigravida | Partial stability | Cumulative adversity | All pregnant people |
| --- | --- | --- | --- | --- | --- |
| **Class proportions** | 0.34 (n = 190) | 0.31 (n = 175) | 0.13 (n = 75) | 0.21 (n = 120) | 1.00 (n = 560) |
| **Socioeconomic characteristics** |  |  |  |  |  |
| **Age category** |  |  |  |  |  |
| Under 25 | 0.00 | 0.03 | 0.13 | 0.17 | 0.06 |
| 25 to 29 | 0.11 | 0.23 | 0.33 | 0.33 | 0.22 |
| 30 to 34 | 0.50 | **0.54** | 0.27 | 0.29 | 0.44 |
| Over 34 | 0.37 | 0.17 | 0.27 | 0.25 | 0.28 |
| **Origin** |  |  |  |  |  |
| The Netherlands | **0.76** | **0.71** | 0.07 | 0.50 | 0.60 |
| One/both parents abroad | 0.11 | 0.11 | 0.40 | 0.25 | 0.18 |
| Abroad | 0.13 | 0.17 | **0.53** | 0.25 | 0.23 |
| **Religion** |  |  |  |  |  |
| No religion | **0.84** | **0.74** | 0.00 | **0.71** | 0.66 |
| Christian/Jewish | 0.13 | 0.23 | 0.20 | 0.17 | 0.19 |
| Muslim | 0.00 | 0.00 | **0.67** | 0.08 | 0.11 |
| Other | 0.03 | 0.03 | 0.07 | 0.04 | 0.04 |
| **Relationship status** |  |  |  |  |  |
| Single/non-cohabiting | 0.00 | 0.03 | 0.00 | 0.33 | 0.09 |
| Cohabiting | 0.42 | **0.51** | 0.20 | **0.54** | 0.45 |
| Married | **0.55** | 0.46 | **0.80** | 0.13 | 0.46 |
| **Single household** |  |  |  |  |  |
| No | **0.97** | **0.89** | **0.93** | 0.46 | 0.82 |
| Yes | 0.03 | 0.11 | 0.07 | **0.54** | 0.18 |
| **Education level** |  |  |  |  |  |
| Low | 0.00 | 0.03 | 0.07 | 0.13 | 0.04 |
| Middle | 0.13 | 0.09 | **0.60** | 0.42 | 0.25 |
| High | **0.87** | **0.89** | 0.33 | 0.46 | 0.71 |
| **Employment** |  |  |  |  |  |
| None | 0.08 | 0.06 | *0.40* | 0.21 | 0.13 |
| Part-time | *0.42* | 0.34 | 0.33 | 0.38 | 0.38 |
| Full-time | 0.50 | **0.63** | 0.27 | 0.42 | 0.49 |
| **Perceived income** |  |  |  |  |  |
| (Very) difficult | 0.00 | 0.00 | 0.13 | *0.17* | 0.05 |
| Coping | 0.11 | 0.09 | 0.33 | *0.50* | 0.21 |
| Comfortable | **0.89** | **0.91** | **0.53** | 0.33 | 0.73 |
| **Income** |  |  |  |  |  |
| Low | 0.08 | 0.09 | **0.53** | **0.63** | 0.26 |
| Middle | **0.63** | **0.57** | 0.33 | 0.33 | 0.51 |
| High | 0.32 | *0.34* | 0.07 | 0.04 | 0.23 |
| **Home ownership** |  |  |  |  |  |
| No | 0.24 | 0.34 | **0.73** | **0.71** | 0.44 |
| Yes | **0.79** | **0.66** | 0.27 | 0.29 | 0.56 |
| **Reproductive characteristics** |  |  |  |  |  |
| **Gestational age at positive test** |  |  |  |  |  |
| Median and under | **0.68** | **0.66** | **0.60** | **0.58** | 0.63 |
| Over median | 0.34 | 0.34 | 0.40 | *0.42* | 0.37 |
| **Gravidity** |  |  |  |  |  |
| Primigravida | 0.00 | **1.00** | 0.33 | 0.38 | 0.45 |
| Multigravida | **1.00** | 0.00 | **0.67** | **0.63** | 0.56 |
| **Previous miscarriage** |  |  |  |  |  |
| No | **0.58** | **1.00** | **0.73** | **0.79** | 0.78 |
| Yes | *0.45* | 0.00 | 0.27 | 0.21 | 0.22 |
| **Previous abortion** |  |  |  |  |  |
| No | **0.84** | **1.00** | **0.93** | **0.75** | 0.89 |
| Yes | 0.16 | 0.00 | 0.07 | *0.25* | 0.11 |
| **Age at sexual debut** |  |  |  |  |  |
| Low | 0.45 | 0.43 | 0.07 | *0.50* | 0.40 |
| Middle | *0.42* | 0.40 | 0.27 | 0.33 | 0.38 |
| High | 0.13 | 0.17 | **0.67** | 0.13 | 0.22 |
| **Negative exposures** |  |  |  |  |  |
| **Worries** |  |  |  |  |  |
| Low | 0.32 | 0.29 | *0.33* | 0.13 | 0.27 |
| Middle | **0.61** | **0.63** | 0.40 | 0.46 | 0.55 |
| High | 0.08 | 0.09 | 0.27 | *0.42* | 0.18 |
| **Recent abuse** |  |  |  |  |  |
| No | **1.00** | **1.00** | **0.93** | **0.88** | 0.96 |
| Yes | 0.00 | 0.00 | 0.07 | *0.13* | 0.04 |
| **Adverse life events** |  |  |  |  |  |
| Zero | *0.08* | 0.03 | 0.07 | 0.00 | 0.04 |
| One to three | **0.76** | **0.74** | **0.67** | 0.29 | 0.64 |
| Four or more | 0.16 | 0.23 | 0.27 | **0.67** | 0.31 |
| **Alcohol use** |  |  |  |  |  |
| Never | 0.08 | 0.14 | **0.87** | 0.25 | 0.25 |
| Stopped during pregnancy | **0.89** | **0.80** | 0.13 | **0.63** | 0.71 |
| Use during pregnancy | 0.03 | 0.06 | 0.00 | *0.13* | 0.05 |
| **Smoking** |  |  |  |  |  |
| Never | **0.87** | **0.89** | **1.00** | 0.46 | 0.80 |
| Stopped during pregnancy | 0.11 | 0.11 | 0.00 | *0.29* | 0.13 |
| Use during pregnancy | 0.03 | 0.00 | 0.00 | *0.21* | 0.06 |
| **Drug use** |  |  |  |  |  |
| No | **0.89** | **0.86** | **1.00** | **0.67** | 0.85 |
| Yes | 0.11 | 0.14 | 0.00 | 0.33 | 0.15 |
| **GP health expenditures** |  |  |  |  |  |
| Low | 0.37 | 0.17 | 0.20 | 0.21 | 0.25 |
| Middle | 0.34 | **0.69** | **0.53** | 0.46 | 0.50 |
| High | 0.29 | 0.11 | 0.27 | 0.38 | 0.25 |
| **Hospital health expenditures** |  |  |  |  |  |
| No costs | 0.37 | 0.46 | 0.33 | 0.42 | 0.39 |
| Lower costs | 0.29 | 0.29 | 0.40 | 0.25 | 0.30 |
| Higher costs | 0.32 | 0.26 | 0.27 | 0.33 | 0.30 |
| **Positive exposures** |  |  |  |  |  |
| **Support** |  |  |  |  |  |
| Low | 0.18 | 0.17 | 0.47 | **0.54** | 0.30 |
| Middle | 0.47 | **0.54** | 0.40 | 0.38 | 0.46 |
| High | 0.34 | 0.29 | 0.13 | 0.08 | 0.23 |
| **Wellbeing** |  |  |  |  |  |
| Low | 0.11 | 0.09 | 0.33 | 0.42 | 0.19 |
| Middle | **0.71** | **0.71** | 0.47 | 0.50 | 0.63 |
| High | 0.21 | 0.20 | 0.20 | 0.13 | 0.18 |
| **Resilience** |  |  |  |  |  |
| Lower | 0.32 | 0.34 | **0.53** | 0.50 | 0.40 |
| Higher | **0.68** | **0.66** | 0.47 | 0.50 | 0.60 |

Note: before calculating proportions, all numbers were rounded to five in accordance with Statistics Netherlands guidelines to prevent disclosure of information about individuals. Hence, proportions may not sum to exactly 1. For each category, the highest class proportion is shown in italics; class proportions >0.5 are shown in bold.

The first (n = 190; 0.34) and second class (n = 175; 0.31) were both characterised by a comfortable perceived income (proportion of 0.89 and 0.91 respectively) and a high education level (0.87, 0.89). Stratified by gravidity, with class 1 being multigravida (1.00) and class 2 being primigravida (1.00), they were respectively labelled *high stability, multigravida* and *high stability, primigravida*. Most were born in the Netherlands (0.76, 0.71) and not religious (0.84, 0.74). Both classes had lower proportions of negative exposures with primarily one to adverse life events (0.76, 0.74) and barely any high worry scores (0.08, 0.09). They generally used alcohol, but stopped during pregnancy (0.89, 0.80) and did not smoke (0.89, 0.86). The *high stability, primigravida* class had higher GP health expenditures than the *high stability, multigravida* class. Further, both classes presented higher proportions of high resilience (0.68, 0.66), and compared to the other two classes, higher wellbeing and support.

The third class (n = 75; 0.13) was particularly characterised by people who had a religion (1.00), being primarily Muslim (0.67). They were often married (0.80) and had a migration background (0.93). Further, they mostly had middle education level (0.60) and had a lower income (0.53), although about half reported a comfortable perceived income (0.53). As for reproductive characteristics, most were multigravida (0.67) and had a higher age at sexual debut (0.67). Negative exposures were low or comparable to class 1 and 2 with regards to substance use, e.g., no smoking was reported, most never used alcohol (0.87), and experienced one to three adverse life events (0.67). With regards to positive exposures, low support (0.47), low wellbeing (0.33), and lower resilience (0.53) stood out, especially compared to class 1 and 2. Class 3 was labelled *partial stability*.

The final class (n = 120; 0.21) was characterised by adversity across multiple dimensions and was thus labelled *cumulative adversity*. Of all classes, this class most often reported their perceived income to be (very) difficult (0.17) or coping (0.50), in line with their proportion of lower income (0.63). They were generally not religious (0.71) and had the highest proportions of being single or in a non-cohabiting relationship (0.33). As for reproductive characteristics, most were multigravida (0.63) and had a lower age at sexual debut (0.50). With regards to negative exposures, the high proportion of four or more adverse life events (0.67) stood out, along with highest proportions of recent abuse (0.13), ever drug use (0.33), and alcohol use (0.13) and smoking (0.21) during pregnancy. Furthermore, this class reports high proportions of low support (0.54), low wellbeing (0.42), and, in comparison with the first two classes, lower resilience (0.50).

### Linear regression

Figure 1 shows the distribution of DA-LMUP scores across all groups, illustrating in pregnancy intention within each class. The figure also indicates differences between classes, especially in the *cumulative adversity* class, with a clearly different distribution pattern. Results from linear regression between class membership and DA-LMUP scores (Table 3) show that when comparing classes to *high stability, multigravida*, two classes had significantly lower pregnancy intentions, with associations remaining after controlling for recruitment location and survey year: *partial stability* (adjusted beta = -0.71 95%CI (-1.31 – -0.11)) and *cumulative adversity* (adjusted beta = -1.81 95%CI (-2.31 – -1.30)). Based on the confidence intervals, there seems to be some overlap between *high stability, primigravida* and *partial stability*, suggesting that the differences between these groups may have been less pronounced, whereas the effect for *cumulative adversity* was distinctly different from all three other classes.

**Figure 1.**
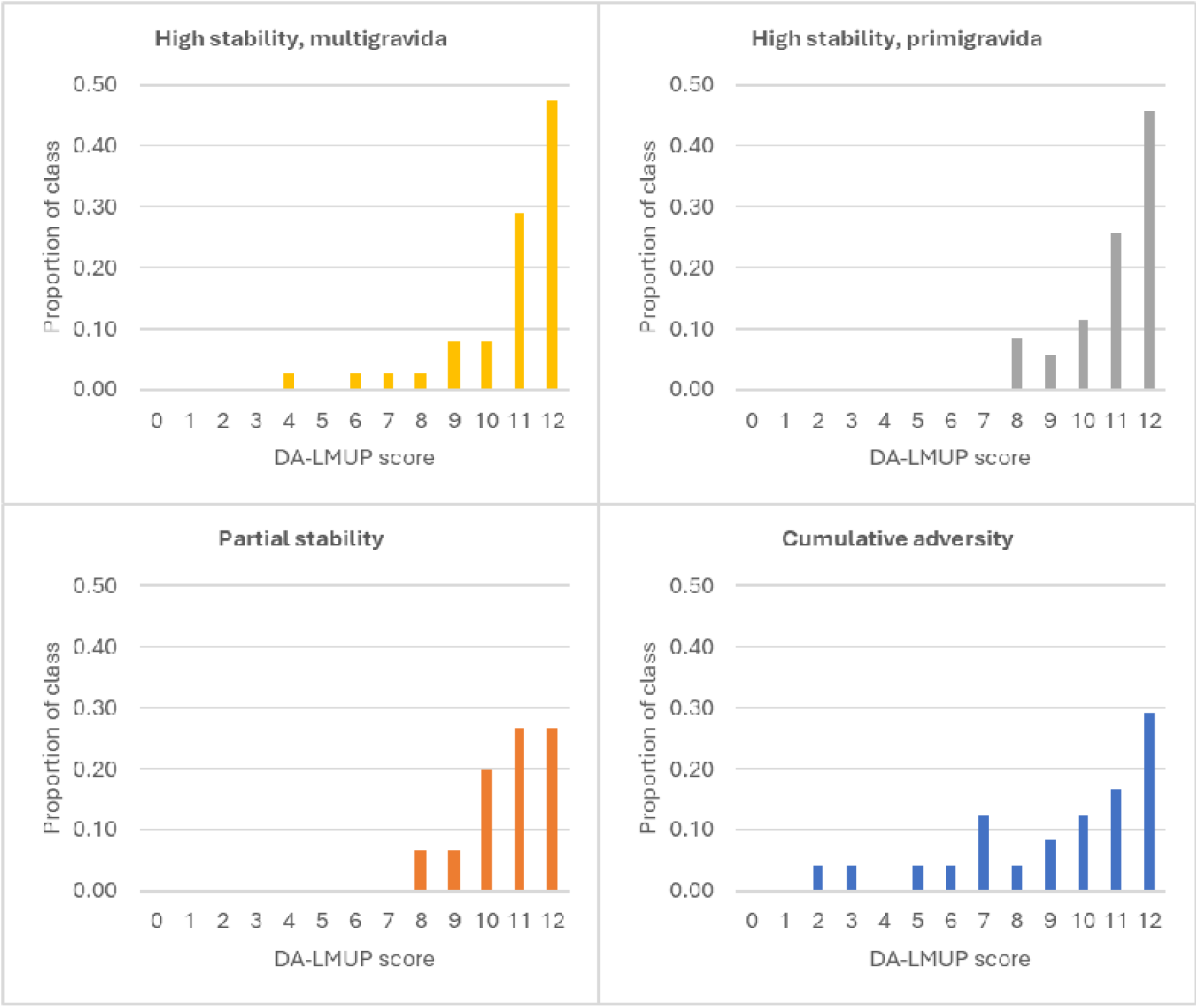
Distribution of pregnancy intention (DA-LMUP) scores for each of the four latent classes.

**Table 3.** Regression analyses on latent class membership and pregnancy intention (DA-LMUP) score.

| Variable | Univariable model<br>(Beta; 95% CI) | P-value | Multivariable model<br>(Beta; 95% CI) | P-value |
| --- | --- | --- | --- | --- |
| High stability, multigravida | Ref. | Ref. | Ref. | Ref. |
| High stability, primigravida | -0.19 (-0.64 – 0.27) | 0.416 | -0.21 (-0.67 – 0.24) | 0.359 |
| Partial stability | **-0.65 (-1.24 – -0.06)** | **0.031** | **-0.71 (-1.31 – -0.11)** | **0.021** |
| Cumulative adversity | **-1.76 (-2.26 – -1.25)** | **<0.001** | **-1.81 (-2.31 – -1.30)** | **<0.001** |

Note: the multivariable model was adjusted for recruitment location, gestational age at survey, and survey year.

## Discussion

In our latent class analysis, we found four distinct classes of pregnant people based on shared combinations of socioeconomic and reproductive characteristics, and positive and negative exposures. Two classes seemed stable, especially socioeconomically and with regards to positive exposures, and differed only on gravidity: all participants classified into the *high stability, multigravida* class had been pregnant before, whereas participants classified into the *high stability, primigravida* class were pregnant for the first time. The two other classes included individuals in more or challenging circumstances. For the *partial stability* class, the instability was primarily socioeconomic. Pregnant people in this class also had lower levels of social support and wellbeing, while they had low levels of substance use. The *cumulative adversity* class reported adversity across different domains, relating to their socioeconomic characteristics, presence of negative exposures including adverse life events and substance use, and lower wellbeing and social support. All classes showed a range of pregnancy intentions. However, compared to the *high stability, multigravida* class, both the *partial stability* class and the *cumulative adversity* class were significantly associated with lower pregnancy intentions. The *cumulative adversity* class also had significantly lower pregnancy intentions compared to the *high stability, primigravida* class. This was not true for the *partial stability* class. Thus, our findings suggest that lower pregnancy intentions are associated with an accumulation of adversity (particularly socioeconomic adversity, adverse life events, and substance use) and low positive exposures (especially in terms of wellbeing and social support).

Compared to earlier work by Molenaar et al.^7^ on combinations of factors relating to vulnerability in pregnancy in the Netherlands, we found similar classes such as to their multidimensional vulnerability class (our cumulative adversity class) and their healthy and socioeconomically stable class was stratified into primigravida and multigravida in our study. Further, our partial stability class might be most comparable to their socioeconomic vulnerability class, a class that showed stability in some areas including low levels of substance use and high proportions of being married while showing less stability regarding income and housing, thus combining both positive and negative exposures.^7^

Regarding pregnancy intentions, our classes with significantly lower pregnancy intentions were similar to some of the identified clusters of women with an unintended pregnancy in a previous study in Rotterdam, the Netherlands.^18^ Two clusters identified in Rotterdam consisted of people with or without a migration background who were younger, single, with lower income and education level, more childhood trauma and lower self-esteem and had similar characteristics as our *cumulative adversity* class.^18^ Another cluster that was more often married, multipara, Islamic, with a migration background, later sexual debut, and lower self-esteem was comparable to the *partial stability* class we found, supporting our findings relating to socioeconomic adversity combined with few positive exposures.^18^

Unlike Molenaar et al.,^7^ who used national registry data prior to pregnancy, we did not identify classes characterised exclusively by high health-care utilisation or psychosocial vulnerability. Similarly, in contrast to Enthoven et al.,^18^ we did not observe a lower pregnancy intention group that was characterised by older age, higher income and education, and a history of depression. This may be attributable to our smaller, actively recruited sample drawn from an urban area in the Netherlands. Although primary factors identified in our study have been associated with unintended pregnancy in prior studies on ‘traditional’ risk factors - being single or in a non-cohabiting relationship, younger or older, lower socioeconomic status, adverse childhood events, periconceptional substance use, and lower support (especially for people with lower income)^8-16^ – our findings, along with the abovementioned studies, show the importance of considering a combination of factors that may influence pregnancy intention or other outcomes.^7,18^

The dominant unintended pregnancy framework in medical care and public health is increasingly put up for debate. The framework has been criticised for approaching pregnancy intentions as a binary construct, and perceiving unintended pregnancy as a problem that needs to be prevented, which in turn might fuel stigmatization.^3,37^ Studies supporting this critiques suggest that pregnancy intentions are not black or white; people do not always have clear ideas about whether they want to become pregnant and previously reported adverse outcomes associated with unintended pregnancy – although small effect sizes – may not be appropriately adjusted for confounding.^38^ Within the prevailing framework, individuals might implicitly be blamed for not preventing pregnancy, especially if in circumstances deemed ‘unfit’ for pregnancy and parenting – for example being young, single, with a low income or a disability. These groups of people are more often addressed by interventions aiming to prevent unintended pregnancy, reinforcing stratified reproduction for example by being encouraged to use long-acting reversible contraception, sometimes through financial incentives or implicit bias.^3,39-41^ However, pregnancy planning might be elusive for people for whom the ‘ideal’ circumstances to have a child might not be within reach and perhaps never will be.^3,37^ Our findings show that people with cumulative adversity and partial stability have lower pregnancy intentions, suggesting systemic inequality rather than individual responsibility. This aligns with a growing body of scientific literature that perceives unintended pregnancy as a symptom of systemic inequality, rather than a marker of reproductive health.^3,37,38,42,43^

### Implications

In light of the critiques on the unintended pregnancy framework, our findings raise the question how best to tailor policy and practice for groups experiencing cumulative adversity while remaining close to individual’s own needs and desires for care and support. One way forward may be to focus on context-sensitive care and support, which starts by asking anyone – not only those labelled as being in ‘vulnerable’ situations –whether they would like to talk about contraception or pregnancy prevention during their visit: the Self-Identified Need for Contraception.^44^ As suggested by Dehlendorf,^38^ this question may be elaborated to include preconception care, focusing on care needs at the moment of the visit rather than intentions for the future.

Furthermore, for individuals experiencing cumulative adversity or partial stability, support needs may extend beyond pregnancy prevention and involve multiple sectors, from health care to state support, requiring strong collaboration across domains. Moniz et al. (2022) have proposed implementing the structural competency framework as an approach to help care providers better address systemic inequality. This might enable them to attend to the structures surrounding the individual person, address needs through referral to (and collaboration with) other services, and advocate for structural change.^40,45,46^

With regards to future research, we encourage scholars to validate these findings and also study parental and child outcomes to establish whether the findings have a clinical impact. Furthermore, future research should investigate how to best meet individuals’ care needs in order to organise care and support in a corresponding manner. Therefore, it might be more relevant to set up a prospective cohort study to study pregnancy intentions (or desires to avoid pregnancy) before people become pregnant.^38,47^ Then, we can truly tailor reproductive policy and practice to preconception, antenatal, and postpartum care needs. Finally, future studies should put more effort into the inclusion of partners and men into pregnancy and reproductive health research, adopting tailored recruitment strategies.

### Strengths and limitations

This study measured pregnancy intentions on a continuum rather than as a binary measure and identified groups characterised by shared combinations of sociodemographic and reproductive characteristics and positive and negative exposures, instead of investigating associations with individual factors. In addition, linking survey data to routine Statistics Netherlands microdata enabled the combination of rich self-reported information with recorded data on for example income, employment, and health-care expenditures.

Nevertheless, several limitations should be considered. First, with regard to the study sample, although we aimed to include both pregnant people and their partners, we did not recruit a sufficient number of partners to conduct the planned analyses. Additionally, we eventually only included people who carried their pregnancy to term and our findings might have been different had we included people who chose abortion. Second, previous research has shown differences in prospective pregnancy intentions between nulliparous, primiparous, and multiparous individuals.^48^ As this distinction could not be made based on our measurements, this may have influenced the results.

Third, we observed some differences between those who did and did not consent to the SN link. People with characteristics resembling the *partial stability* class were therefore underrepresented, which may limit internal validity, as our analyses relied on the subsample that provided consent. Considering this and given the convenience sampling approach, we cannot draw conclusions about the external validity of the observed associations, although the characteristics of the identified groups are mostly in line with previous findings in the Netherlands.^7,18^ Moreover, as this study was conducted within the context of a high income welfare state with relatively extensive access to reproductive health care and social welfare, the extent to which the findings are generalisable to settings with different health care systems and social welfare arrangements remains uncertain.

Fourth, class labels were assigned to facilitate interpretation but should be used with caution, as people are placed into a class based on probability; no participant is fully representative of their assigned class. Finally, due to the observational study design, causal inferences cannot be made and risk of residual confounding from unmeasured factors – such as earlier contraceptive experiences, positive childhood experiences, or experiences of racism – remains.^3,49,50^

## Conclusion

This study identified four latent classes of pregnant people, showing that lower pregnancy intentions were associated with cumulative adversity as well as with partial stability supporting literature framing unintended pregnancy as a symptom of structural inequalities. We suggest shifting away from labelling individuals as vulnerable towards asking what care or support they need related to the level of pregnancy intent. A structural competency framework may help providers to consider the circumstances shaping people’s lives and to collaborate across domains to better address needs from people with cumulative adversity. Future research should prospectively study reproductive intentions prior to conception and better include partners to tailor policy and practice to reproductive preferences.

## Data Availability

Research data are not shared in accordance with Statistics Netherlands guidelines to prevent disclosure of information about individuals.

## References

1. Nelson HD, Darney BG, Ahrens K, et al. Associations of Unintended Pregnancy With Maternal and Infant Health Outcomes. JAMA 2022; 328(17): 1714.

2. Aiken AR, Westhoff CL, Trussell J, Castano PM. Comparison of a Timing-Based Measure of Unintended Pregnancy and the London Measure of Unplanned Pregnancy. Perspect Sex Reprod Health 2016; 48(3): 139–46.

3. Auerbach SL, Coleman-Minahan K, Alspaugh A, Aztlan EA, Stern L, Simmonds K. Critiquing the unintended pregnancy framework. Journal of Midwifery & Women’s Health 2023; 68(2): 170–8.

4. Gipson JD, Koenig MA, Hindin MJ. The effects of unintended pregnancy on Infant, Child, and Parental Health: A Review of the Literature. Stud Fam Plann 2008; 39(1): 18–38.

5. Klerman LV. The intendedness of pregnancy: A concept in transition. Matern Child Health J 2000; 4(3): 155–62.

6. Barrett G, Smith SC, Wellings K. Conceptualisation, development, and evaluation of a measure of unplanned pregnancy. J Epidemiol Community Health 2004; 58(5): 426–33.

7. Molenaar JM, van der Meer L, Bertens LCM, et al. Defining vulnerability subgroups among pregnant women using pre-pregnancy information: a latent class analysis. Eur J Public Health 2023; 33(1): 25–34.

8. Goossens J, Van Den Branden Y, Van Der Sluys L, et al. The prevalence of unplanned pregnancy ending in birth, associated factors, and health outcomes. Human Reproduction 2016; 31(12): 2821–33.

9. Finer LB, Zolna MR. Declines in Unintended Pregnancy in the United States, 2008–2011. N Engl J Med 2016; 374(9): 843-52.

10. Hall JA, Barrett G, Copas A, Stephenson J. London Measure of Unplanned Pregnancy: guidance for its use as an outcome measure. Patient Relat Outcome Meas 2017; 8: 43–56.

11. Feld H, Barnhart S, Wiggins AT, Ashford K. Social support reduces the risk of unintended pregnancy in a low-income population. Public Health Nursing 2021; 38(5): 801–9.

12. Li X, Yang B, Nie L, Ren Z, Sun Y, Liu J. Association between adverse childhood experiences and unintended pregnancy: A systematic review and meta-analysis. Child Abuse & Neglect 2025; 169: 107642.

13. Lundsberg LS, Pensak MJ, Gariepy AM. Is Periconceptional Substance Use Associated with Unintended Pregnancy? Women’s Health Reports 2020; 1(1): 17–25.

14. Shafique S, Umer A, Innes KE, Rudisill TM, Fang W, Cottrell L. Preconception Substance Use and Risk of Unintended Pregnancy: Pregnancy Risk Assessment Monitoring System 2016-17. Journal of addiction medicine 2022; 16(3): 278–85.

15. Iseyemi AB, Zhao QM, McNicholas CD, Peipert JF. Socioeconomic Status As a Risk Factor for Unintended Pregnancy in the Contraceptive CHOICE Project. Obstetrics & Gynecology 2017; 130(3): 609–15.

16. Barton K, Redshaw M, Quigley MA, Carson C. Unplanned pregnancy and subsequent psychological distress in partnered women: a cross-sectional study of the role of relationship quality and wider social support. BMC Pregnancy and Childbirth 2017; 17(1).

17. Carlander A, Hultstrand JN, Reuterwall I, Jonsson M, Tydén T, Kullinger M. Unplanned pregnancy and the association with maternal health and pregnancy outcomes: A Swedish cohort study. PLOS ONE 2023; 18(5): e0286052.

18. Enthoven CA, El Marroun H, Koopman-Verhoeff ME, et al. Clustering of characteristics associated with unplanned pregnancies: the generation R study. BMC Public Health 2022; 22(1).

19. Ardesch FH, Meulendijk MC, Kist JM, et al. The introduction of a data-driven population health management approach in the Netherlands since 2019: The Extramural LUMC Academic Network data infrastructure. Health Policy 2023; 132: 104769.

20. National Academies of Sciences E, Medicine, Health, et al. In: Negussie Y, Geller A, DeVoe JE, eds. Vibrant and Healthy Kids: Aligning Science, Practice, and Policy to Advance Health Equity. Washington (DC): National Academies Press (US) Copyright 2019 by the National Academy of Sciences. All rights reserved.; 2019.

21. Molenaar JM, Leung KY, Van Der Meer L, Klein PPF, Struijs JN, Kiefte-De Jong JC. Predicting population-level vulnerability among pregnant women using routinely collected data and the added relevance of self-reported data. European Journal of Public Health 2024; 34(6): 1210–7.

22. De Groot N, Bonsel GJ, Birnie E, Valentine NB. Towards a universal concept of vulnerability: Broadening the evidence from the elderly to perinatal health using a Delphi approach. PLOS ONE 2019; 14(2): e0212633.

23. Colciago E, Merazzi B, Panzeri M, Fumagalli S, Nespoli A. Women’s vulnerability within the childbearing continuum: A scoping review. European Journal of Midwifery 2020; 4(May).

24. Van der Meer L, Ernst H, Blanchette L, Steegers E. Een kwetsbare zwangere, wat is dat eigenlijk. Medisch Contact 2020; 22: 34–6.

25. Green JM, Kafetsios K, Statham HE, Snowdon CM. Factor Structure, Validity and Reliability of the Cambridge Worry Scale in a Pregnant Population. Journal of Health Psychology 2003; 8(6): 753–64.

26. Kliem S, Mößle T, Rehbein F, Hellmann DF, Zenger M, Brähler E. A brief form of the Perceived Social Support Questionnaire (F-SozU) was developed, validated, and standardized. Journal of Clinical Epidemiology 2015; 68(5): 551–62.

27. Butjosa A, Gómez-Benito J, Myin-Germeys I, et al. Development and validation of the Questionnaire of Stressful Life Events (QSLE). Journal of Psychiatric Research 2017; 95: 213–23.

28. Connor KM, Davidson JRT. Development of a new resilience scale: The Connor-Davidson Resilience Scale (CD-RISC). Depression and Anxiety 2003; 18(2): 76–82.

29. McFarlane J, Parker B, Soeken K, Bullock L. Assessing for Abuse During Pregnancy: Severity and Frequency of Injuries and Associated Entry Into Prenatal Care. Journal of the American Medical Association 1992; 267: 3176–8.

30. Stewart-Brown S, Tennant A, Tennant R, Platt S, Parkinson J, Weich S. Internal construct validity of the Warwick-Edinburgh Mental Well-being Scale (WEMWBS): a Rasch analysis using data from the Scottish Health Education Population Survey. Health and Quality of Life Outcomes 2009; 7(1): 15.

31. Sprenger M, Beumer WY, Van Ditzhuijzen J, Kiefte-De Jong JC. Pregnancy intentions in the Netherlands: An evaluation of a multidimensional and continuous construct. medRxiv; 2025.

32. Van Buuren S. Flexible Imputation of Missing Data. 2nd Edition ed. New York: Chapman and Hall/CRC; 2018.

33. Collins LM. Latent class and latent transition analysis : with applications in the social, behavioral, and health sciences: Hoboken, NJ : Wiley & Sons; 2010.

34. Linzer DA, Lewis JB. poLCA: An R Package for Polytomous Variable Latent Class Analysis. Journal of Statistical Software 2011; 42(10): 1–29.

35. Nylund KL, Asparouhov T, Muthén BO. Deciding on the Number of Classes in Latent Class Analysis and Growth Mixture Modeling: A Monte Carlo Simulation Study. Structural Equation Modeling: A Multidisciplinary Journal 2007; 14(4): 535–69.

36. Jung T, Wickrama KAS. An Introduction to Latent Class Growth Analysis and Growth Mixture Modeling. Social and Personality Psychology Compass 2008; 2(1): 302–17.

37. Macleod CI. Public reproductive health and ‘unintended’ pregnancies: introducing the construct ‘supportability’. Journal of Public Health 2016; 38(3): e384–e91.

38. Dehlendorf C, Perry JC, Borrero S, Callegari L, Fuentes L, Perritt J. Meeting people’s pregnancy prevention needs: Let’s not force people to state an “Intention”. Contraception 2024; 135: 110400.

39. Eeckhaut MCW, Hara Y. Reproductive Oppression Enters the Twenty-First Century: Pressure to Use Long-Acting Reversible Contraception (LARC) in the Context of “LARC First”. Socius: Sociological Research for a Dynamic World 2023; 9.

40. Moniz MH, Spector-Bagdady K, Perritt JB, et al. Balancing enhanced contraceptive access with risk of reproductive injustice: A United States comparative case study. Contraception 2022; 113: 88–94.

41. Colen S. "Like a Mother to Them": Stratified Reproduction and West Indian Childcare Workers and Employers in New York. In: Ginsburg FD, Rapp R, eds. Conceiving the New World Order: The Global Politics of Reproduction. Berkeley, CA: University of California Press; 1995.

42. Beumer WY, Roseboom TJ, Van Ditzhuijzen J. What contributes to pregnancy intendedness? Insights from the Dutch BluePrInt study using a conceptual hierarchical model. Cold Spring Harbor Laboratory; 2025.

43. Doria CM, Liddell JL, Buscaglia A, Buxbaum L, Gliko S. Exploring Contraceptive Experiences of Abortion-Fund Clients in the Rocky Mountain Region of the United States from a Reproductive Justice Lens. Womens Reprod Health (Phila) 2024; 11(3): 491–508.

44. Person-Centered Reproductive Health Program. SINC Implementation Guide. 2025. https://pcrhp.ucsf.edu/sinc-implementation-guide.

45. Metzl JM, Hansen H. Structural competency: Theorizing a new medical engagement with stigma and inequality. Social Science & Medicine 2014; 103: 126–33.

46. Downey MM, Gómez AM. Structural Competency and Reproductive Health. AMA J Ethics 2018; 20(3): 211–23.

47. Rocca CH, Ralph LJ, Wilson M, Gould H, Foster DG. Psychometric Evaluation of an Instrument to Measure Prospective Pregnancy Preferences. Medical Care 2019; 57(2): 152–8.

48. Bullington BW, Muñoz I, Boscardin WJ, Rocca CH. Pregnancy Preferences and Incident Pregnancy in the US. JAMA Network Open 2025; 8(10): e2536697.

49. Swiatlo A, Curtis S, Gottfredson N, Halpern C, Tumlinson K, Lich KH. Contraceptive Behavior Dynamics and Unintended Pregnancy A Latent Transition Analysis. Demography 2023; 60(4): 1089–113.

50. Merrick JS, Narayan AJ, Atzl VM, Harris WW, Lieberman AF. Type versus timing of adverse and benevolent childhood experiences for pregnant women’s psychological and reproductive health. Children and Youth Services Review 2020; 114: 105056.

