## Supplementary Tables for "Clustering of characteristics associated with level of pregnancy intention: a latent class analysis of an urban cross-sectional sample in the Netherlands"

### Supplementary Table 1. Description of measures

| **Measure** | **Categories** | **Data source** | **Additional information** |
| --- | --- | --- | --- |
| **Sociodemographic characteristics** | | | |
| Age | Categorised based on commonly used cut-offs: <25 years; 25 to 29 years; 30 to 34 years; >34 years. | RISE UP survey |  |
| Gender | Male; Female; Other | RISE UP survey |  |
| Origin | Categorised based on SN standards: participant and parents born in the Netherlands; participant born in the Netherlands, one or both parents born abroad; participant born abroad. | RISE UP survey |  |
| Religion | Not religious; Christian or Jewish; Muslim; Hindu, Buddhist, or other. | RISE UP survey |  |
| Marital status | Married; cohabiting relationship; non-cohabiting relationship; single.  In analyses: non-cohabiting relationship and single merged. | RISE UP survey |  |
| Single household | No; yes. | SN microdata | Data available until 2023, data taken from reference year July-June. |
| Education level | Low; middle; high. | SN microdata | Data available until 2023, data taken from reference year July-June. |
| Employment | None; part-time; full-time | SN microdata | Data available until 2024, data taken from reference year July-June. |
| Perceived income | Very difficult or difficult; coping; comfortable. | RISE UP survey |  |
| Income percentile | Skewed distribution, categorised based on quartiles: low (Q1), middle (Q2-3), and high (Q4). | SN microdata | Data on household income available until 2023, data taken from reference year July-June. |
| Home ownership | No; yes. | SN microdata | Data available until 2023, data taken from reference year July-June. |
| **Reproductive characteristics** | | | |
| Gestational age at positive test | In weeks. Extremely skewed distribution, categorised based on median, median included in largest group: earlier; later | RISE UP survey |  |
| Gestational age at survey | In weeks. Treated as continuous variable. | RISE UP survey |  |
| Gravidity | Primigravida; multigravida | RISE UP survey |  |
| Previous miscarriage | No; yes. | RISE UP survey |  |
| Previous abortion | No; yes. | RISE UP survey |  |
| Age at sexual debut | Skewed distribution, categorised based on quartiles: low (Q1), middle (Q2-3), and high (Q4). | RISE UP survey |  |
| **Negative exposures** | | | |
| Worries | Skewed distribution, categorised based on quartiles: low (Q1), middle (Q2-3), and high (Q4). | RISE UP survey | Cambridge Worry Scale (CWS),^[1]^ first eight questions: average score. |
| Recent abuse | No; yes. | RISE UP survey | Physical, sexual, or emotional abuse in the year before or during pregnancy |
| Adverse life events | Categorised based on commonly used cut-offs. Zero events; one to three events; four or more events. | RISE UP survey | Selection of question of Questionnaire of Stressful Life Events (QSLE)^[2]^ based on representativeness scores. |
| Alcohol use | No alcohol use before pregnancy; stopped alcohol use during pregnancy; alcohol use during pregnancy. | RISE UP survey |  |
| Smoking | No smoking before pregnancy; stopped smoking during pregnancy; smoking during pregnancy. | RISE UP survey |  |
| Drug use | No (no drugs before pregnancy); Yes (stopped drug use during pregnancy or drug use during pregnancy). | RISE UP survey |  |
| GP expenditures | Skewed distribution, categorised based on quartiles: low (Q1), middle (Q2-3), and high (Q4). | SN microdata | Data available until 2022, data taken from year before survey or, if pregnancy in that year, two years before survey. |
| Hospital expenditures | Extremely skewed distribution including large proportion with no expenditures, further categorised based on the median of nonzero expenditures (pregnant people: 399.20, partners: 383.59): no costs; lower; higher. | SN microdata | Data available until 2022, data taken from year before survey or, if pregnancy in that year, two years before survey. |
| **Positive exposures** | | | |
| Support | Skewed distribution, categorised based on quartiles: low (Q1), middle (Q2-3), and high (Q4). | RISE UP survey | Brief form of the Perceived Social Support Questionnaire (F-SozU K6):^[3]^ sum score. |
| Wellbeing | Normally distributed, categorised based on mean plus or minus 1 standard deviation: low; middle; high. | RISE UP survey | Short Warwick-Edinburgh Mental Wellbeing Scale (SWEMWBS):^[4]^ transformed to metric sum score based on guidelines. |
| Resilience | Extremely skewed distribution, categorised based on median, median included in largest group: lower; higher. | RISE UP survey | Connor-Davidson Resilience Scale (CD-RISC2):^[5]^ sum score. |
| **Outcome** | | | |
| Pregnancy intention | Treated as continuous variable. | RISE UP survey | Dutch Adaptation of the London Measure of Unplanned Pregnancy (DA-LMUP).^[6]^ |
| **Recruitment characteristics** | | | |
| Recruitment location | Youth and Family Centre; Primary care midwife; Online; Other. | RISE UP informed consent |  |
| Survey year | 2022; 2023; 2024/2025 | RISE UP survey | Year in which survey was completed.  Note: SN microdata is reported for calendar years (January-December). Therefore, SN microdata was extracted for the previous year if the survey was completed in January-June and for the survey year if the survey was completed in July-December – e.g., when a survey was completed in June 2022, the reference year was 2021 for characteristics and exposures extracted from SN microdata, and when a survey was completed in July 2022, the reference year was 2022. |

### Supplementary Table 2. Characteristics of participants with and without consent for SN link, n (%) unless noted otherwise.

|  | **Pregnant people** | |  | **Partners** |  |  |
| --- | --- | --- | --- | --- | --- | --- |
|  | **No consent for SN** | **Consent for SN** | **All pregnant people** | **No consent for SN** | **Consent for SN** | **All partners** |
| **Measure** | (N=95) | (N=560) | (N=655) | (N=20) | (N=125) | (N=145) |
| **Socioeconomic characteristics** |  |  |  |  |  |  |
| **Age – mean (SD)** | 30.3 (4.7) | 31.8 (4.5) | 31.6 (4.6) | 32.1 (5.5) | 33 (5.4) | 32.9 (5.4) |
| Under 25 | 10 (12) | 35 (6) | 45 (7) | 5 (14) | 5 (3) | 5 (5) |
| 25-29 | 30 (32) | 125 (22) | 155 (24) | 0 (9) | 25 (20) | 25 (19) |
| 30-34 | 35 (36) | 245 (44) | 280 (43) | 10 (46) | 55 (43) | 65 (43) |
| 35 and over | 20 (20) | 155 (28) | 175 (26) | 5 (32) | 40 (33) | 50 (33) |
| **Gender** |  |  |  |  |  |  |
| Man | 0 (1) | 0 (0) | 0 (0) | 20 (100) | 115 (95) | 140 (96) |
| Woman | 95 (99) | 555 (99) | 650 (99) | 0 (0) | 5 (5) | 5 (4) |
| Other | 0 (0) | 5 (1) | 5 (1) | 0 (0) | 0 (0) | 0 (0) |
| Missing | 0 (0) | 0 (0) | 0 (0) | 0 (0) | 0 (0) | 0 (0) |
| **Origin** |  |  |  |  |  |  |
| The Netherlands | 40 (44) | 335 (59) | 375 (57) | 15 (59) | 85 (68) | 95 (66) |
| Abroad: one or both parents | 35 (36) | 100 (18) | 130 (20) | 5 (23) | 20 (17) | 25 (18) |
| Abroad: self | 20 (20) | 130 (23) | 145 (22) | 0 (9) | 15 (14) | 20 (13) |
| Missing | 0 (0) | 0 (0) | 0 (0) | 0 (9) | 0 (2) | 5 (3) |
| **Religion** |  |  |  |  |  |  |
| No religion | 40 (45) | 365 (65) | 405 (62) | 10 (41) | 80 (67) | 90 (63) |
| Christian/Jewish | 25 (25) | 95 (17) | 120 (18) | 5 (32) | 20 (15) | 25 (18) |
| Muslim | 20 (23) | 60 (10) | 80 (12) | 0 (9) | 10 (9) | 15 (9) |
| Other | 5 (4) | 25 (5) | 30 (5) | 0 (9) | 5 (5) | 10 (6) |
| Missing | 5 (3) | 15 (3) | 20 (3) | 0 (9) | 5 (4) | 5 (5) |
| **Relationship status** |  |  |  |  |  |  |
| Single | 5 (3) | 30 (5) | 30 (5) | 0 (5) | 0 (0) | 0 (1) |
| Non-cohabiting relationship | 5 (4) | 20 (4) | 25 (4) | 0 (9) | 0 (1) | 5 (2) |
| Cohabiting relationship | 35 (36) | 250 (45) | 285 (43) | 10 (41) | 50 (42) | 60 (41) |
| Married | 55 (56) | 260 (46) | 315 (48) | 10 (46) | 70 (57) | 80 (55) |
| Missing | 0 (0) | 0 (0) | 0 (0) | 0 (0) | 0 (1) | 0 (1) |
| **Perceived income** |  |  |  |  |  |  |
| (Very) difficult | 5 (5) | 30 (5) | 35 (5) | 0 (0) | 5 (6) | 5 (5) |
| Coping | 30 (31) | 115 (21) | 145 (22) | 5 (18) | 30 (23) | 30 (22) |
| Comfortable | 55 (60) | 405 (72) | 460 (71) | 15 (77) | 80 (67) | 100 (68) |
| Missing | 5 (4) | 10 (2) | 15 (2) | 0 (5) | 5 (5) | 5 (5) |
| **Reproductive characteristics** |  |  |  |  |  |  |
| **Gestational age at positive test – median (IQR)** | 4 (4-5) | 4 (4-5) | 4 (4-5) | 4 (3-6.25) | 4 (3-6) | 4 (3-6) |
| Median and under | 50 (54) | 355 (63) | 405 (62) | 10 (50) | 75 (61) | 85 (59) |
| Over median | 40 (45) | 205 (37) | 250 (38) | 10 (41) | 45 (36) | 55 (37) |
| Missing | 0 (1) | 0 (0) | 0 (0) | 0 (9) | 5 (3) | 5 (4) |
| **Gestational age at survey – median (IQR)** | 24 (18.25-28) | 24 (15-27) | 24 (15-27) | 24 (14-28) | 25 (14-28) | 25 (14-28) |
| **Gravidity** |  |  |  |  |  |  |
| Primigravida | 40 (43) | 245 (44) | 285 (44) | 10 (36) | 70 (55) | 75 (52) |
| Multigravida | 50 (53) | 310 (55) | 360 (55) | 10 (55) | 50 (39) | 60 (41) |
| Missing | 5 (4) | 5 (1) | 5 (1) | 0 (9) | 5 (6) | 10 (6) |
| **Previous miscarriage** |  |  |  |  |  |  |
| No | 60 (64) | 425 (76) | 485 (74) | 15 (73) | 95 (76) | 110 (75) |
| Yes | 30 (30) | 125 (22) | 150 (23) | 5 (18) | 25 (19) | 25 (19) |
| Missing | 5 (6) | 10 (2) | 20 (3) | 0 (9) | 5 (6) | 10 (6) |
| **Previous abortion** |  |  |  |  |  |  |
| No | 80 (84) | 495 (88) | 575 (88) | 15 (77) | 110 (90) | 130 (88) |
| Yes | 10 (10) | 60 (11) | 70 (11) | 5 (14) | 5 (4) | 10 (6) |
| Missing | 5 (6) | 5 (1) | 10 (1) | 0 (9) | 5 (6) | 10 (6) |
| **Age at sexual debut – median (IQR)** | 18 (16-21) | 17 (16-19) | 17 (16-19) | 18 (16-21.75) | 18 (16-20) | 18 (16-20) |
| Low | 30 (33) | 220 (39) | 250 (38) | 10 (41) | 40 (34) | 50 (35) |
| Middle | 30 (32) | 205 (37) | 235 (36) | 5 (23) | 55 (43) | 60 (40) |
| High | 30 (31) | 120 (22) | 150 (23) | 10 (36) | 25 (21) | 35 (23) |
| Missing | 5 (4) | 15 (2) | 15 (3) | 0 (0) | 0 (2) | 0 (1) |
| **Negative exposures** |  |  |  |  |  |  |
| **Worries – median (IQR)** | 0.38 (0.13-0.84 | 0.38 (0.13-0.88) | 0.38 (0.13-0.88) | 0.25 (0.13-0.53) | 0.50 (0.25-0.75) | 0.38 (0.13-0.75) |
| Low | 30 (30) | 145 (26) | 175 (26) | 5 (32) | 30 (24) | 35 (26) |
| Middle | 45 (48) | 310 (55) | 355 (54) | 10 (50) | 65 (51) | 75 (51) |
| High | 20 (22) | 100 (18) | 120 (18) | 0 (9) | 30 (23) | 30 (21) |
| Missing | 0 (0) | 5 (1) | 5 (1) | 0 (9) | 0 (2) | 5 (3) |
| **Recent abuse** |  |  |  |  |  |  |
| No | 90 (98) | 530 (94) | 620 (95) | 20 (91) | 115 (92) | 135 (92) |
| Yes | 0 (1) | 20 (3) | 20 (3) | 0 (0) | 5 (6) | 5 (5) |
| Missing | 0 (1) | 15 (2) | 15 (2) | 0 (9) | 5 (2) | 5 (3) |
| **Adverse life events – median** (**IQR)** | 2 (1-3) | 3 (1-4) | 2 (1-4) | 3 (1-4) | 3 (1-4) | 3 (1-4) |
| Zero | 5 (6) | 30 (5) | 35 (6) | 5 (18) | 10 (8) | 15 (10) |
| One to three | 65 (69) | 360 (64) | 425 (65) | 10 (50) | 65 (52) | 75 (52) |
| Four or more | 25 (25) | 170 (30) | 195 (30) | 5 (32) | 50 (40) | 55 (39) |
| **Alcohol use** |  |  |  |  |  |  |
| Never | 45 (46) | 135 (24) | 180 (27) | 5 (27) | 30 (23) | 35 (23) |
| Stopped during pregnancy | 45 (47) | 385 (69) | 430 (66) | 5 (14) | 20 (15) | 20 (15) |
| Had alcohol during pregnancy | 5 (6) | 30 (5) | 35 (5) | 10 (55) | 70 (59) | 85 (58) |
| Missing | 0 (1) | 10 (2) | 15 (2) | 0 (5) | 5 (3) | 5 (3) |
| **Smoking** |  |  |  |  |  |  |
| Never | 75 (82) | 450 (80) | 525 (81) | 15 (68) | 100 (81) | 115 (79) |
| Stopped during pregnancy | 10 (12) | 75 (13) | 85 (13) | 0 (9) | 5 (6) | 10 (6) |
| Had alcohol during pregnancy | 5 (6) | 30 (6) | 40 (6) | 5 (18) | 15 (13) | 20 (14) |
| Missing | 0 (0) | 5 (1) | 5 (1) | 0 (5) | 0 (1) | 0 (1) |
| **Drug use** |  |  |  |  |  |  |
| No | 85 (92) | 475 (85) | 560 (86) | 15 (77) | 95 (78) | 115 (78) |
| Yes | 10 (9) | 80 (14) | 90 (14) | 5 (14) | 25 (20) | 30 (19) |
| Missing | 0 (0) | 5 (1) | 5 (1) | 0 (9) | 0 (2) | 5 (3) |
| **Positive exposures** |  |  |  |  |  |  |
| **Wellbeing – mean (SD)** | 25.5 (4.4) | 25.6 (3.9) | 25.6 (4) | 27.2 (5) | 26.2 (3.8) | 26.3 (4) |
| Low | 15 (18) | 100 (18) | 115 (18) | 5 (18) | 15 (11) | 15 (12) |
| Middle | 55 (61) | 380 (68) | 435 (67) | 10 (50) | 85 (68) | 95 (65) |
| High | 10 (13) | 65 (12) | 80 (12) | 5 (32) | 20 (15) | 25 (17) |
| Missing | 10 (9) | 15 (3) | 25 (4) | 0 (0) | 10 (7) | 10 (6) |
| **Resilience – median (IQR)** | 6 (5-7) | 7 (6-8) | 7 (6-8) | 6 (5-8) | 7 (6-8) | 7 (6-8) |
| Under median | 50 (55) | 240 (43) | 295 (45) | 10 (55) | 50 (40) | 60 (42) |
| Median and above | 40 (45) | 320 (57) | 360 (55) | 10 (46) | 75 (60) | 85 (58) |
| **Support – median (IQR)** | 26 (23-29) | 27 (24-29) | 27 (24-29) | 25.5 (24.25-28.75) | 26 (24-29) | 26 (24-29) |
| Low | 40 (42) | 175 (32) | 215 (33) | 5 (27) | 45 (36) | 50 (35) |
| Middle | 35 (39) | 255 (46) | 295 (45) | 15 (59) | 55 (46) | 70 (48) |
| High | 20 (19) | 125 (23) | 145 (22) | 5 (14) | 25 (19) | 25 (18) |
| **Recruitment characteristics** |  |  |  |  |  |  |
| **Recruitment location** |  |  |  |  |  |  |
| Youth and Family Centre | 35 (38) | 225 (40) | 260 (40) | 5 (18) | 30 (23) | 30 (22) |
| Primary care midwife | 20 (21) | 140 (25) | 160 (25) | 10 (36) | 50 (42) | 60 (41) |
| Online | 20 (19) | 130 (23) | 145 (22) | 0 (5) | 10 (9) | 10 (8) |
| Other | 20 (21) | 65 (12) | 85 (13) | 10 (41) | 30 (26) | 40 (28) |
| **Survey year** |  |  |  |  |  |  |
| 2022 | 15 (14) | 120 (21) | 130 (21) | 5 (14) | 30 (24) | 30 (22) |
| 2023 | 50 (52) | 265 (47) | 315 (48) | 10 (50) | 50 (40) | 60 (41) |
| 2024/2025 | 30 (34) | 180 (32) | 210 (32) | 10 (36) | 45 (37) | 55 (37) |
| **Outcome** |  |  |  |  |  |  |
| **Pregnancy intention – median (IQR)** | 11 (10-12) | 11 (10-12) | 11 (10-12) | 9 (8-10) | 9 (8-10) | 9 (8-10) |

Note: table reports imputed data. Numbers rounded to five and percentages presented as integer numbers in accordance with Statistics Netherlands guidelines to prevent disclosure of information about individuals. Hence, percentages may not sum to exactly 100.

### Supplementary Table 3. Fit indices of latent class models with increasing number of classes

| Number of classes | Size-adjusted Bayesian Information Criterion (aBIC) | Entropy |
| --- | --- | --- |
| 1 | 23110.3 | NA |
| 2 | 22204.3 | 0.822 |
| 3 | 21951.3 | 0.859 |
| 4 | 21826.3 | 0.927 |
| 5 | 21749.5 | 0.872 |
| 6 | 21732.5 | 0.865 |
| 7 | 21744.0 | 0.844 |
| 8 | 21747.5 | 0.731 |
| 9 | 21790.6 | 0.794 |
| 10 | 21794.1 | 0.866 |

1. Green, J. M., Kafetsios, K., Statham, H. E., & Snowdon, C. M. (2003). Factor Structure, Validity and Reliability of the Cambridge Worry Scale in a Pregnant Population. *Journal of Health Psychology*, *8*(6), 753-764.
2. Butjosa, A., Gómez-Benito, J., Myin-Germeys, I., Barajas, A., Baños, I., Usall, J., Grau, N., Granell, L., Sola, A., Carlson, J., Dolz, M., Sánchez, B., Haro, J. M., Araya, S., Arranz, B., Arteaga, M., Asensio, R., Autonell, J., Baños, I., ... Ochoa, S. (2017). Development and validation of the Questionnaire of Stressful Life Events (QSLE). *Journal of Psychiatric Research*, *95*, 213-223. https://doi.org/https://doi.org/10.1016/j.jpsychires.2017.08.016
3. Kliem, S., Mößle, T., Rehbein, F., Hellmann, D. F., Zenger, M., & Brähler, E. (2015). A brief form of the Perceived Social Support Questionnaire (F-SozU) was developed, validated, and standardized. *Journal of Clinical Epidemiology*, *68*(5), 551-562. https://doi.org/https://doi.org/10.1016/j.jclinepi.2014.11.003
4. Ng Fat, L., Scholes, S., Boniface, S., Mindell, J., & Stewart-Brown, S. (2017). Evaluating and establishing national norms for mental wellbeing using the short Warwick–Edinburgh Mental Well-being Scale (SWEMWBS): findings from the Health Survey for England. *Quality of Life Research*, *26*(5), 1129-1144. https://doi.org/10.1007/s11136-016-1454-8
5. Vaishnavi, S., Connor, K., & Davidson, J. R. T. (2007). An abbreviated version of the Connor-Davidson Resilience Scale (CD-RISC), the CD-RISC2: psychometric properties and applications in psychopharmacological trials. *Psychiatry research*, *152*(2-3), 293-297. https://doi.org/10.1016/j.psychres.2007.01.006
6. Sprenger, M., Beumer, W. Y., Van Ditzhuijzen, J., & Kiefte-De Jong, J. C. (2025). Pregnancy intentions in the Netherlands: An evaluation of a multidimensional and continuous construct. *medRxiv*. https://www.medrxiv.org/content/10.1101/2023.11.13.23298453v3
